# A Novel Dataset for Nuclei and Tissue Segmentation in Melanoma with baseline nuclei segmentation and tissue segmentation benchmarks

**DOI:** 10.1101/2024.10.07.24315039

**Authors:** Mark Schuiveling, Hong Liu, Daniel Eek, Gerben E. Breimer, Karijn P.M. Suijkerbuijk, Willeke A.M. Blokx, Mitko Veta

## Abstract

**Background:** Melanoma is an aggressive form of skin cancer in which tumor-infiltrating lymphocytes (TILs) are a biomarker for recurrence and treatment response. Manual TIL assessment is prone to interobserver variability, and current deep learning models are not publicly accessible or have low performance. Deep learning models, however, have the potential of consistent spatial evaluation of TILs and other immune cell subsets with the potential of improved prognostic and predictive value. To make the development of these models possible, we created the Panoptic Segmentation of nUclei and tissue in advanced MelanomA (PUMA) dataset and assessed the performance of several state-of-the-art deep learning models. In addition, we show how to improve model performance further by using heuristic post-processing in which nuclei classes are updated based on their tissue localization.

**Results:** The PUMA dataset includes 155 primary and 155 metastatic melanoma H&E stained regions of interest with nuclei and tissue annotations from a single melanoma referral institution. The Hover-NeXt model, trained on the PUMA dataset, demonstrated the best performance for lymphocyte detection, approaching human interobserver agreement. In addition, heuristic post-processing of deep learning models improve the detection of non-common classes, such as epithelial nuclei.

**Conclusion:** The PUMA dataset is the first melanoma specific dataset that can be used to develop melanoma-specific nuclei and tissue segmentation models. These models can, in turn, be used for prognostic and predictive biomarker development. Incorporating tissue and nuclei segmentation is a step towards improved deep learning nuclei segmentation performance. We will use this dataset to organize the PUMA challenge in which the goal is to further improve model performance.

## Background

Melanoma is an aggressive form of skin cancer with increasing incidence [1]. Primary melanoma is treated with surgical excision, whereas advanced, metastasized melanoma is most commonly treated with immune checkpoint inhibition therapy, a form of cancer immunotherapy. However, half of the advanced melanoma patients do not respond to this treatment, which is costly and potentially toxic [2–5].

Previous studies showed that tumor infiltrating lymphocytes (TILs) in hematoxylin & eosin (H&E) stained slides of metastatic melanoma before the start of treatment are associated with a higher chance of response to immune checkpoint inhibition and an increase in survival [6,7]. In addition, TILs are associated with reduced recurrence rates in primary melanoma [8]. Therefore, assessment of TILs can serve as a prognostic biomarker in melanoma treatment.

Currently, TILs are scored manually, either by estimating a stromal percentage or through using a multitier system like the Clark score, which categorizes TILs as absent, non-brisk, or brisk [6,9,10]. However, substantial interobserver variability exists among pathologists [6,11,12]. A deep learning-based assessment could result in a more precise, consistent and fine-grained assessment of TILs.

To date, two deep learning-based models have been used to evaluate TILs in melanoma histopathology. The first model, NN192, uses watershed segmentation followed by a fully connected neural network [7]; however, this older, suboptimal technique results in lower performance [13]. The second model, LUNIT’s Scope IO pan tumor model, is not melanoma specific and not publicly available [14,15]. Other publicly available models, such as Hover-Net pretrained on the PanNuke dataset that includes skin samples can be used to detect TILs in melanoma, but have suboptimal performance. This is due to the model not being melanoma-specific [16,17]. As a result, misclassifications occur because melanoma cells can resemble other cell types, such as stroma or lymphocytes [13,18,19].

**Figure 1.**
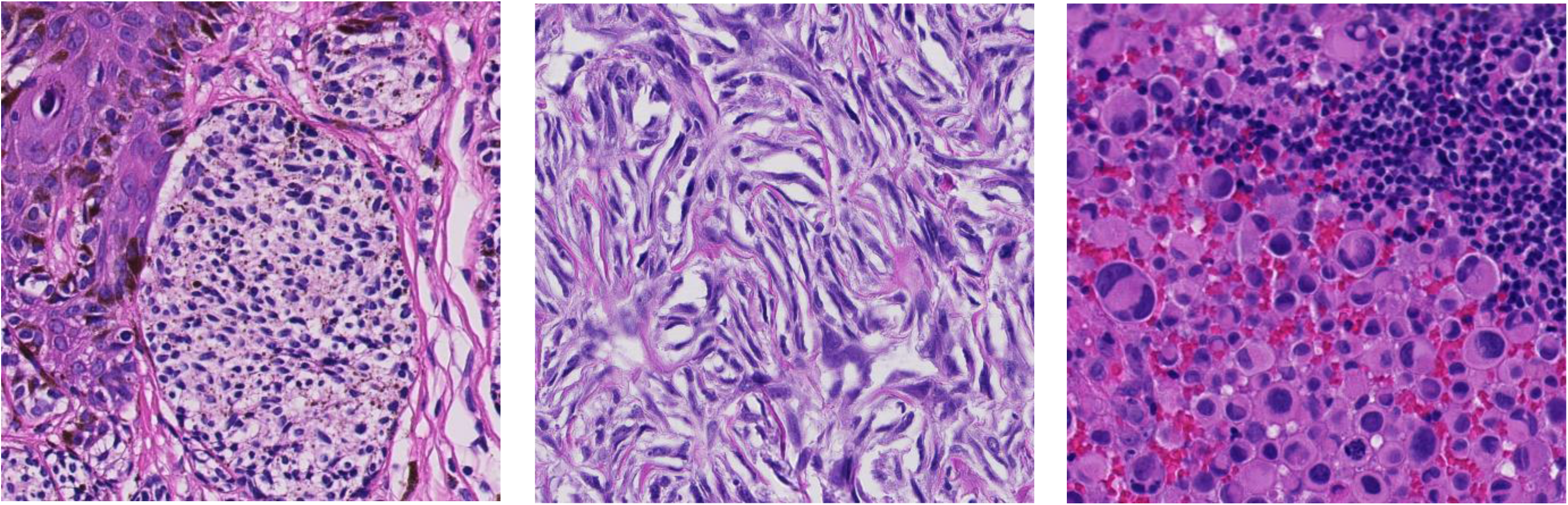
Examples of melanoma histopathology appearances: a lymphocyte-like morphology on the left, a stromal growth pattern in the center, and a tumor with large, variable nuclei on the right.

Next to the presence of TILs, the localization of TILs is of importance. In breast cancer and non-small cell lung cancer, TILs located within the tumor or intra-tumoral stroma regions, rather than in necrotic tissue, are predictive of outcomes [20,21]. No public dataset with tissue annotations or public model capable of segmenting melanoma tumor and necrotic tissue regions exists at this moment.

Furthermore, other immune cell subsets might also have prognostic implications. For example, neutrophils are associated with an increased chance of primary melanoma metastasizing, and B cell presence is associated with response to immune checkpoint inhibition therapy in melanoma [22,23]. The 2020 MoNuSAC challenge showed that it is possible to segment immune cell subsets in H&E stained histopathology images [24].

Consequently, there is a need for a deep learning model capable of segmenting nuclei of tumor cells and different immune cell subsets in H&E slides of melanoma. In addition, such a model should be capable of segmenting tissue areas which can be used for nuclei localization. To address these needs, we created the Panoptic Segmentation of nUclei and tissue in advanced MelanomA (PUMA) dataset. In this paper, we describe the methodology used to create the dataset. Furthermore, we provide nuclei instance segmentation and tissue semantic segmentation benchmarks as well as a first step in improving nuclei segmentation due to the integration of a tissue and nuclei segmentation.

### Data description

The PUMA dataset consists of regions of interest (ROIs) with nuclei and tissue annotations. ROIs originate from H&E stained histological slides of melanoma specimens. The dataset’s goal is to facilitate the development of deep learning models capable of segmenting nuclei and tissue. To stimulate the use of the dataset and create novel deep learning models, the dataset will also be used for a medical image analysis challenge hosted on the grand-challenge.org platform. The models created with the dataset and the PUMA challenge, in turn, can be used for prognostic biomarker generation in melanoma treatment.

The dataset consists of 155 primary and 155 metastatic melanoma manually selected ROIs, scanned at 40× magnification (0.22 µm/px) with a resolution of 1024 × 1024 pixels. For these ROIs, annotations of both tissue and nuclei are supplied, as well as a context ROI of 5120 × 5120 pixels centered around the ROI. Annotations were created by a medical expert (author M.S.) and checked and corrected by a dermatopathologist (author W.B.). All cases were digitized in a large melanoma referral center, however 76 cases are revisions or consultations originating from other treatment hospitals. Annotations are in the .GeoJSON format, making annotations easily visualizable with the opensource pathology image viewer Qupath [25,26].

A total of 103 primary and 103 metastatic ROI have been made publicly available [27]. The remaining 104 annotated ROIs are kept private as an independent test set to be used in the PUMA challenge. The public set consists of 97 429 nuclei in 103 primary melanoma ROIs and 103 metastatic melanoma ROIs. The test set consists of 50 490 nuclei in 52 primary and 52 metastatic ROIs. The distribution of nuclei types and metastatic sample location is visualized in Figure 2.

**Figure 2.**
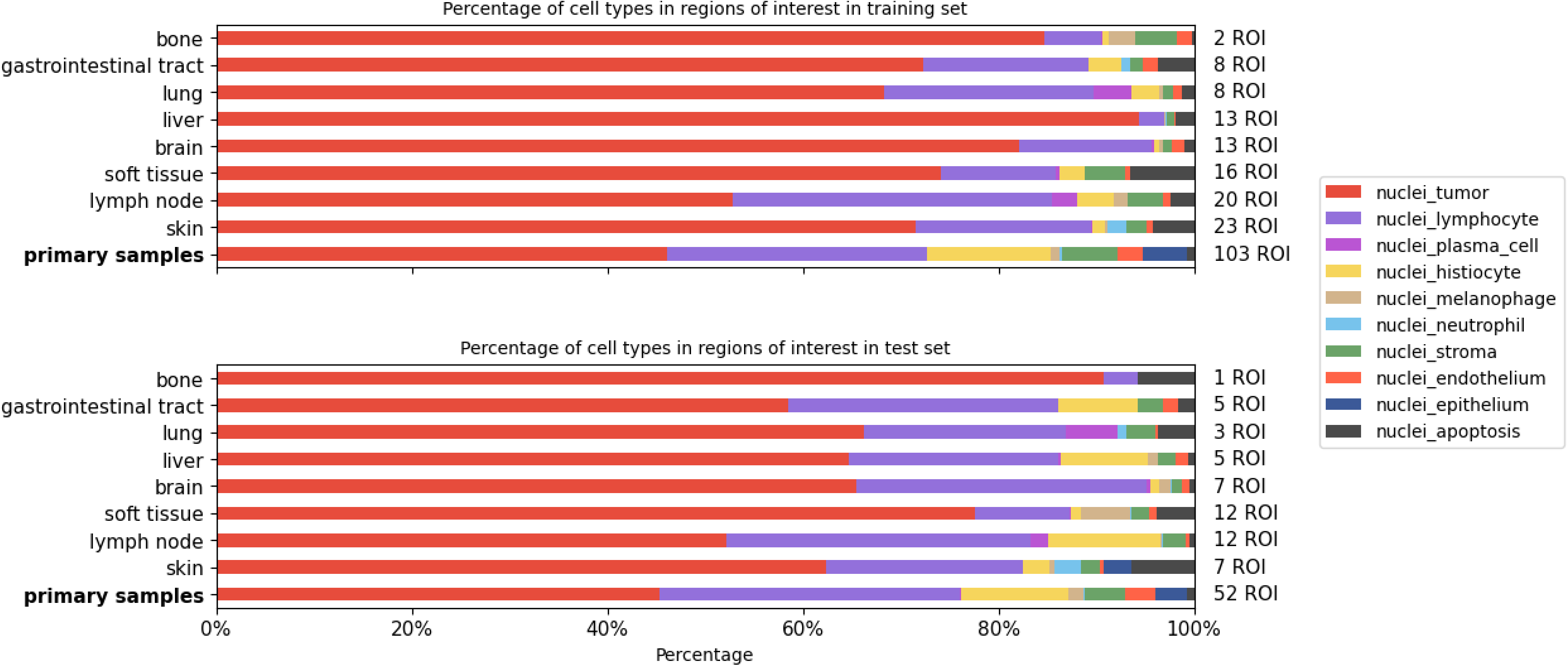
Distribution of nuclei and sampled tissue in the PUMA dataset for primary melanoma regions of interest and metastatic melanoma regions of interest stratified according to metastasis location.

As we published previously, more lymphocytes are present in primary samples than in metastatic samples [6]. The most common metastatic lesion sites are lymph nodes and skin metastases. In all samples, lymphocytes and tumor nuclei form the majority of nuclei.

The tissue distribution for the training and test sets is visualized in Figure 3. In primary samples, more tumoral stroma is present compared to metastatic samples. The epidermis and necrotic area are underrepresented in both datasets.

**Figure 3.**
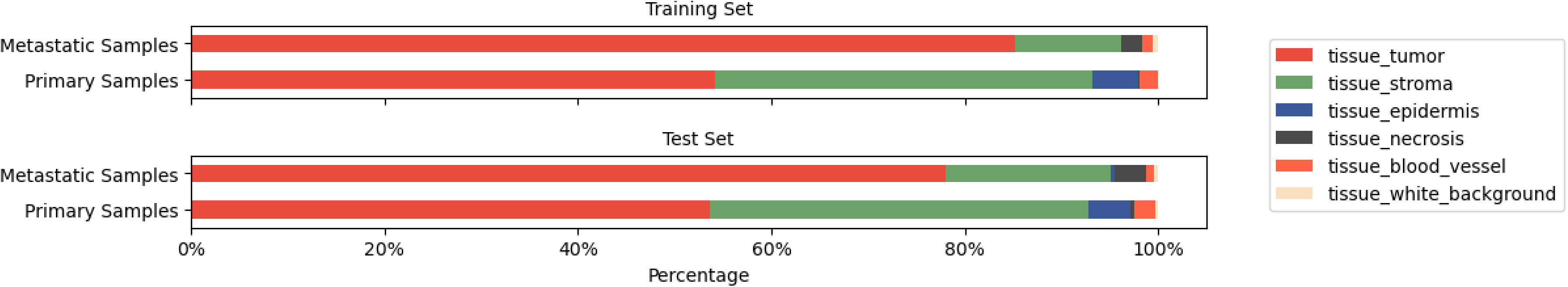
Distribution of tissue class area in the PUMA dataset.

A detailed description of the dataset creation process can be found in the methods section.

#### Analyses

To set a baseline for the PUMA dataset, we performed four experiments. The first experiment was semantic segmentation of the tumor, stroma, epidermis, blood vessel, and necrotic tissue classes. The second experiment was nuclei segmentation with three nuclei classes: tumor, lymphocyte, and other. For the third experiment, we performed nuclei segmentation with all nuclei classes: tumor, lymphocyte, plasma cell, histiocyte, melanophage, neutrophil, stroma, endothelium, epithelium, and apoptosis. Finally, we show an incorporation of tissue predictions to update the nuclei predictions as a form of heuristic post-processing.

We used 5-fold cross validation while training and report the results of inference on the 94 ROIs of the final independent test set used in the PUMA challenge. In addition, we report the results of the inter- and intraobserver agreement as performed on 12 random samples.

#### Tissue segmentation

In the first experiment, semantic segmentation of tissue was performed with a nnU-Net model and a Mask2Former model with the backbone replaced by the UNI pathology foundation model [28–30]. The goal of this analysis was to assess to what extent semantic segmentation of different tissue classes is possible with state-of-the-art segmentation models and to evaluate how this correlates with intra- and interobserver agreement. For evaluation of segmentation, the Dice score was computed for each class per sample and averaged across all samples (referred to as average Dice). Additionally, a Dice score was calculated per class on a concatenated sample. To create this sample, all images were combined along one axis, resulting in a single large image with the width of one image and a length equal to the number of images × the height of one image. This is referred to as the micro Dice.

Dice scores are visualized in Table 1 and a visualization of the segmentation is presented in Figure 4. The nnU-Net model achieved the highest overall Dice scores but could not recognize necrosis in the dataset. While Mask2Former Dice scores were lower, it could detect a small part of the necrosis area. When compared to the DICE score of intra- and interobserver agreement, the Dice scores for both models were low in the stroma, epidermis, blood vessel, and necrosis classes.

**Figure 4.**
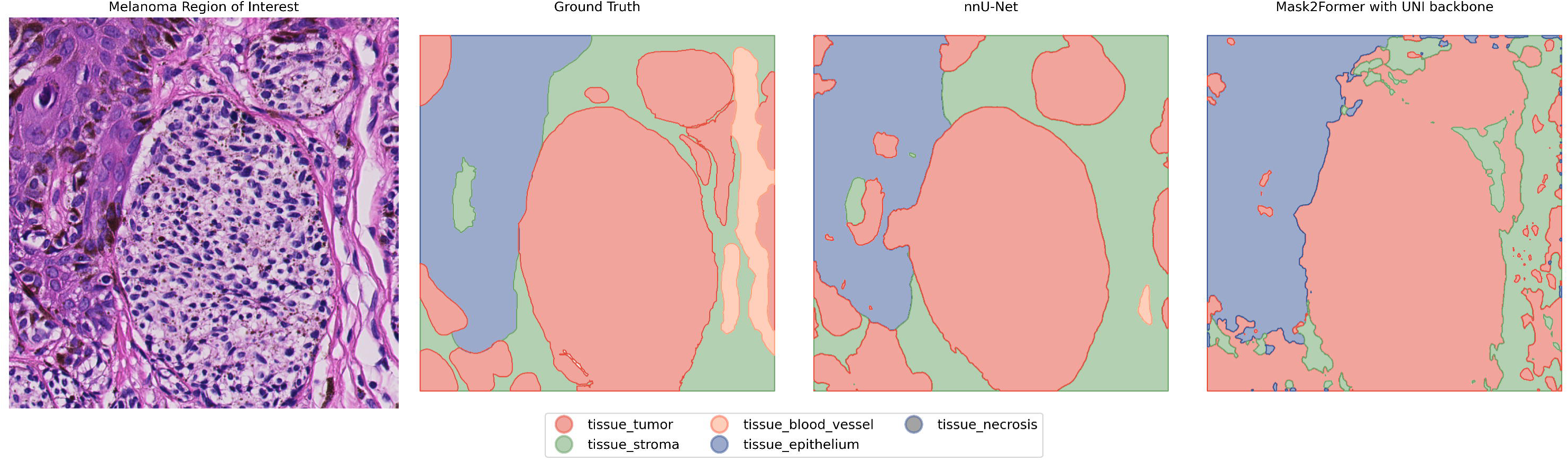
The visual result of semantic segmentation of tissue with the ground truth, the result of the nnU-Net model, and the Mask2Former model.

**Table 1.** Average and micro average Dice scores of semantic segmentation of tissue over all samples. Results are displayed for a nnU-Net model and a Mask2Former model with the backbone replaced by the UNI pathology foundation model. For comparison, DICE scores of intra- and interobsever agreement are displayed.

|  | Class | Tumor<br>[95% CI] | Stroma<br>[95% CI] | Epidermis<br>[95% CI] | Blood Vessel<br>[95% CI] | Necrosis<br>[95% CI] | Average<br>[95% CI] |
| --- | --- | --- | --- | --- | --- | --- | --- |
| nnU-Net | Average | 0.87 [0.83 - 0.90] | 0.59 [0.52 - 0.68] | 0.91 [0.86 - 0.96] | 0.54 [0.45 - 0.63] | 0.93 [0.87 - 0.98] | 0.77 [0.73 - 0.80] |
|  | Micro Average | 0.91 [0.88 - 0.94] | 0.78 [0.71 - 0.84] | 0.69 [0.42 - 0.86] | 0.35 [0.23 - 0.47] | 0.01 [0.00 - 0.04] | 0.55 [0.54 - 0.55] |
| Mask2-Former<br>(UNI backbone) | Average | 0.81 [0.76 - 0.86] | 0.47 [0.39 - 0.55] | 0.90 [0.85 - 0.96] | 0.46 [0.36 - 0.57] | 0.92 [0.86 - 0.97] | 0.71 [0.68 - 0.75] |
|  | Micro Average | 0.86 [0.82 - 0.89] | 0.62 [0.52 - 0.71] | 0.63 [0.34 - 0.84] | 0.01 [0.00 - 0.02] | 0.09 [0.00 - 0.23] | 0.44 [0.43 - 0.44] |
| Intra-observer | Average | 0.96 [0.93 - 0.99] | 0.89 [0.72 - 0.99] | 1.00 [0.99 - 1.0] | 0.86 [0.75 - 0.95] | 0.97 [0.92 - 1.0] | 0.94 [0.90 - 0.97] |
|  | Micro Average | 0.98 [0.95 - 0.99] | 0.94 [0.90 - 0.97] | 0.98 [0.98 - 1.0] | 0.68 [0.52 - 0.79] | 0.91 [0.70 - 1.0] | 0.90 [0.90 - 0.91] |
| Inter-observer | Average | 0.97 [0.95 - 0.98] | 0.89 [0.73 - 0.99] | 1.00 [0.99 - 1.00] | 0.72 [0.50 - 0.9] | 0.97 [0.91 - 1.0] | 0.91 [0.87 - 0.95] |
|  | Micro Average | 0.98 [0.97 - 0.99] | 0.94 [0.89 - 0.97] | 0.97 [0.96 - 1.0] | 0.73 [0.59 - 0.83] | 0.90 [0.67 - 1.0] | 0.90 [0.90 - 0.90] |

#### Segmentation of three nuclei classes

In the second experiment, nuclei segmentation was performed for three classes: tumor nuclei, lymphocytes (including plasma nuclei), and other. The goal of the analysis was to evaluate the usability of existing models in skin and/or melanoma histopathology, compare them with models trained on our dataset, and assess how this correlates with intra- and interobserver agreement. For this experiment we compared the NN192 model, Hover-Net and Hover-NeXt trained on the PanNuke dataset (a pan-tissue dataset that includes skin tissue) and Hover-Net and Hover-NeXt trained on our dataset. Among all evaluated models, the Hover-NeXt model trained on the PUMA dataset using three classes as input for training, demonstrated the highest performance with a F_1_ score for lymphocytes close to the intra- and interobserver agreement.

The NN192 model, a melanoma specific model trained to recognize lymphocytes, showed the lowest performance. The Hover-Net and Hover-NeXt models trained on the PanNuke dataset were more successful in detecting lymphocytes. However, the PanNuke-trained Hover-Net model tended to misclassify tumor nuclei as lymphocytes. Models trained on the PUMA dataset, logically, had better performance on evaluation on the test dataset. Results are displayed in Table 2, and an example of the model predictions is shown in Figure 5.

**Figure 5.**
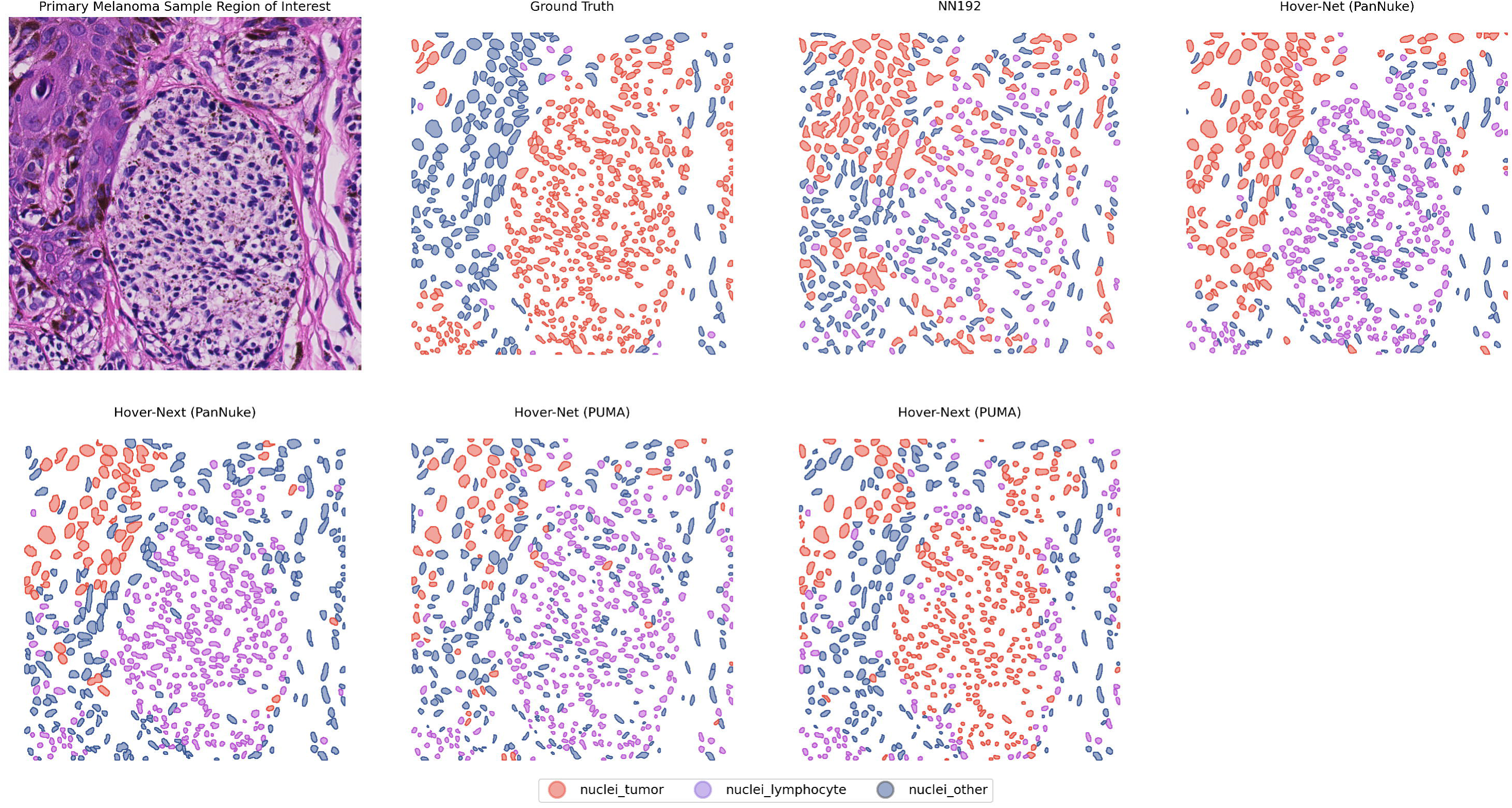
Visual results of segmentation of tumor nuclei, lymphocytes and other nuclei. Results are shown for the NN192 model, the Hover-Net and Hover-NeXt model trained on the PanNuke dataset and the Hover-Net and Hover-NeXt model trained on the PUMA dataset.

**Table 2.** F_1_ scores for segmentation of tumor, lymphocyte, and other nuclei categories. Results are shown for intra- and interobserver agreement, the NN192 model, Hover-Net and Hover-NeXt trained on the PanNuke dataset and Hover-Net and Hover-NeXt trained on the PUMA dataset.

|  | **Tumor**<br>[95% CI] | **Lymphocyte**<br>[95% CI] | **Other**<br>[95% CI] | **Micro $F_1$**<br>[95% CI] | **Average $F_1$**<br>[95% CI] |
| --- | --- | --- | --- | --- | --- |
| **Intraobserver agreement** | 0.92 [0.9 - 0.94] | 0.85 [0.8 - 0.89] | 0.82 [0.77 - 0.86] | 0.89 [0.89 - 0.89] | 0.86 [0.85 - 0.87] |
| **Interobserver agreement** | 0.89 [0.87 - 0.91] | 0.83 [0.79 - 0.88] | 0.76 [0.71 - 0.81] | 0.85 [0.85 - 0.85] | 0.83 [0.82 - 0.84] |
| **NN192** | 0.59 [0.59 - 0.6] | 0.11 [0.10 - 0.11] | 0.13 [0.12 - 0.14] | 0.46 [0.46 - 0.46] | 0.28 [0.27 - 0.28] |
| **Hover-Net (PanNuke)** | 0.74 [0.74 - 0.75] | 0.57 [0.55 - 0.58] | 0.37 [0.36 - 0.38] | 0.64 [0.64 - 0.64] | 0.56 [0.56 - 0.56] |
| **Hover-NeXt (PanNuke)** | 0.68 [0.67 - 0.68] | 0.68 [0.67 - 0.70] | 0.38 [0.37 - 0.39] | 0.61 [0.61 - 0.61] | 0.58 [0.58 - 0.58] |
| **Hover-Net (PUMA)** | 0.74 [0.73 - 0.74] | 0.69 [0.68 - 0.69] | 0.41 [0.41 - 0.42] | 0.66 [0.66 - 0.66] | 0.61 [0.61 - 0.61] |
| **Hover-NeXt (PUMA)** | 0.81 [0.81 - 0.81] | 0.80 [0.79 - 0.81] | 0.50 [0.49 - 0.51] | 0.76 [0.76 - 0.76] | 0.70 [0.70 - 0.71] |

#### Classification of all nuclei classes

In the third experiment, segmentation was performed for all nuclei classes within the PUMA dataset using Hover-Net and Hover-NeXt. In this analysis, we evaluated to what extent Hover-Net and Hover-NeXt are able to correctly segment all nuclei classes present in the dataset. In addition, this analysis aimed at establishing intra- and interobserver agreement for all classes.

Both Hover-Net and Hover-Next showed low performance in terms of segmentation of all non-common classes (Table 3). In addition, the F_1_ scores for Tumor and Lymphocyte were lower when compared to the same models trained with three nuclei classes. In the visual representation (Figure 5) the decreased capacity of the Hover-NeXt model of segmenting difficult cases is clearly visible. Inter- and intraobserver F_1_ scores are lower due to disagreement in the classification of plasma cells, melanophages, stroma nuclei, and endothelium. In the 12 randomly selected samples used to assess intra- and interobserver agreement, plasma cells and melanophages had a low incidence; 2 plasma cells, and 14 melanophages were present (Table 4). Stroma, endothelium, and histiocytes had higher incidence but were still subject to substantial disagreement.

**Table 3.**
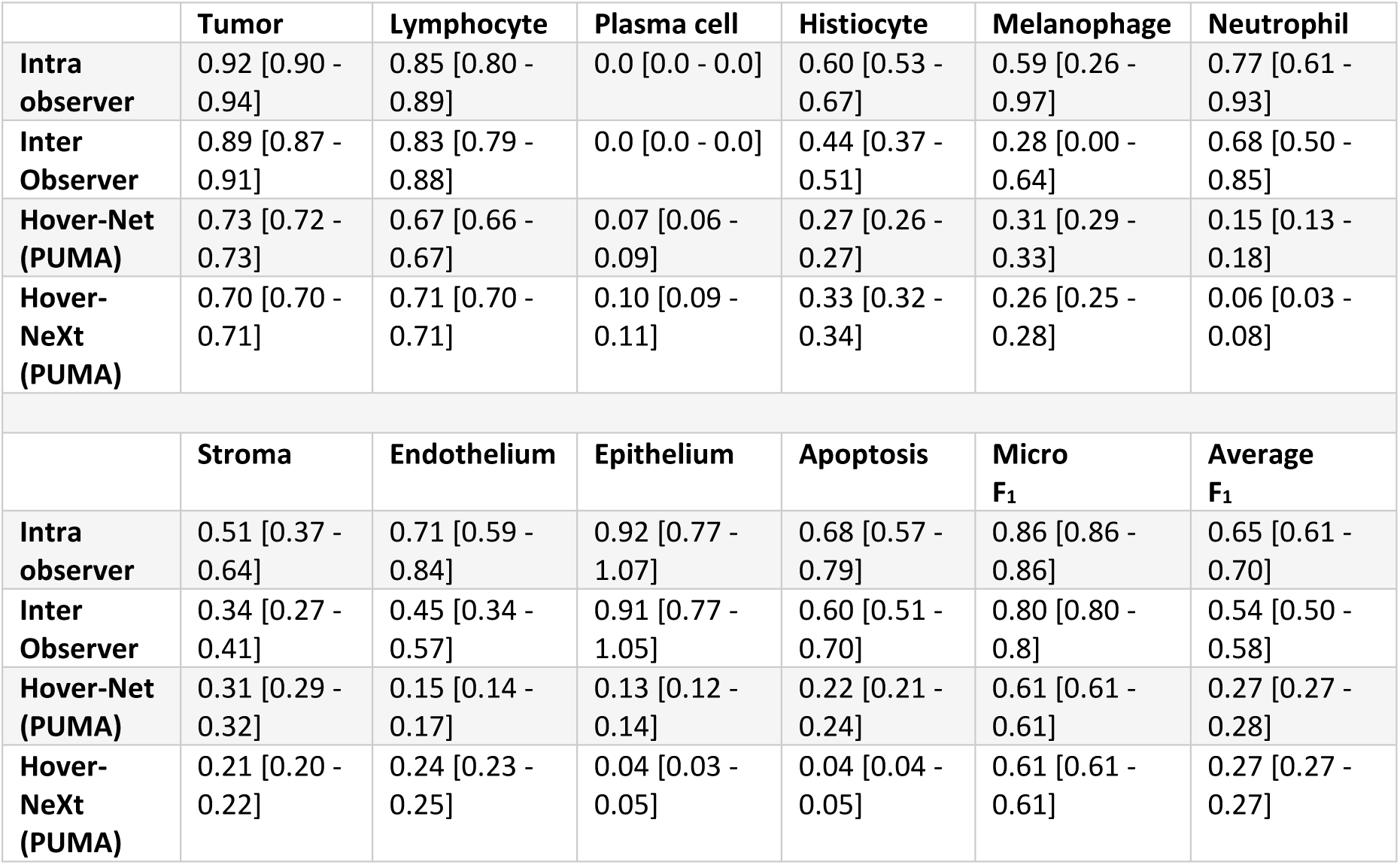
F_1_ scores for intra- and interobserver agreement and model segmentation in all classes.

|  | **Tumor** | **Lymphocyte** | **Plasma cell** | **Histiocyte** | **Melanophage** | **Neutrophil** |
| --- | --- | --- | --- | --- | --- | --- |
| **Intra observer** | 0.92 [0.90 - 0.94] | 0.85 [0.80 - 0.89] | 0.0 [0.0 - 0.0] | 0.60 [0.53 - 0.67] | 0.59 [0.26 - 0.97] | 0.77 [0.61 - 0.93] |
| **Inter Observer** | 0.89 [0.87 - 0.91] | 0.83 [0.79 - 0.88] | 0.0 [0.0 - 0.0] | 0.44 [0.37 - 0.51] | 0.28 [0.00 - 0.64] | 0.68 [0.50 - 0.85] |
| **Hover-Net (PUMA)** | 0.73 [0.72 - 0.73] | 0.67 [0.66 - 0.67] | 0.07 [0.06 - 0.09] | 0.27 [0.26 - 0.27] | 0.31 [0.29 - 0.33] | 0.15 [0.13 - 0.18] |
| **Hover-NeXt (PUMA)** | 0.70 [0.70 - 0.71] | 0.71 [0.70 - 0.71] | 0.10 [0.09 - 0.11] | 0.33 [0.32 - 0.34] | 0.26 [0.25 - 0.28] | 0.06 [0.03 - 0.08] |
|  | **Stroma** | **Endothelium** | **Epithelium** | **Apoptosis** | **Micro $F_1$** | **Average $F_1$** |
| **Intra observer** | 0.51 [0.37 - 0.64] | 0.71 [0.59 - 0.84] | 0.92 [0.77 - 1.07] | 0.68 [0.57 - 0.79] | 0.86 [0.86 - 0.86] | 0.65 [0.61 - 0.70] |
| **Inter Observer** | 0.34 [0.27 - 0.41] | 0.45 [0.34 - 0.57] | 0.91 [0.77 - 1.05] | 0.60 [0.51 - 0.70] | 0.80 [0.80 - 0.8] | 0.54 [0.50 - 0.58] |
| **Hover-Net (PUMA)** | 0.31 [0.29 - 0.32] | 0.15 [0.14 - 0.17] | 0.13 [0.12 - 0.14] | 0.22 [0.21 - 0.24] | 0.61 [0.61 - 0.61] | 0.27 [0.27 - 0.28] |
| **Hover-NeXt (PUMA)** | 0.21 [0.20 - 0.22] | 0.24 [0.23 - 0.25] | 0.04 [0.03 - 0.05] | 0.04 [0.04 - 0.05] | 0.61 [0.61 - 0.61] | 0.27 [0.27 - 0.27] |

**Table 4.**
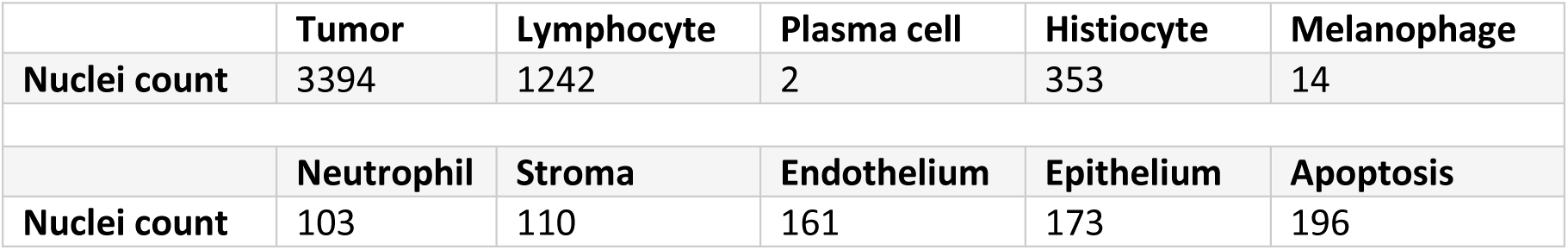
Nuclei count in intra and inter-observer ground truth dataset consisting out of 12 samples.

|  | **Tumor** | **Lymphocyte** | **Plasma cell** | **Histiocyte** | **Melanophages** |
| --- | --- | --- | --- | --- | --- |
| **Nuclei count** | 3394 | 1242 | 2 | 353 | 14 |
|  | **Neutrophil** | **Stroma** | **Endothelium** | **Epithelium** | **Apoptosis** |
| **Nuclei count** | 103 | 110 | 161 | 173 | 196 |

**Figure 6.**
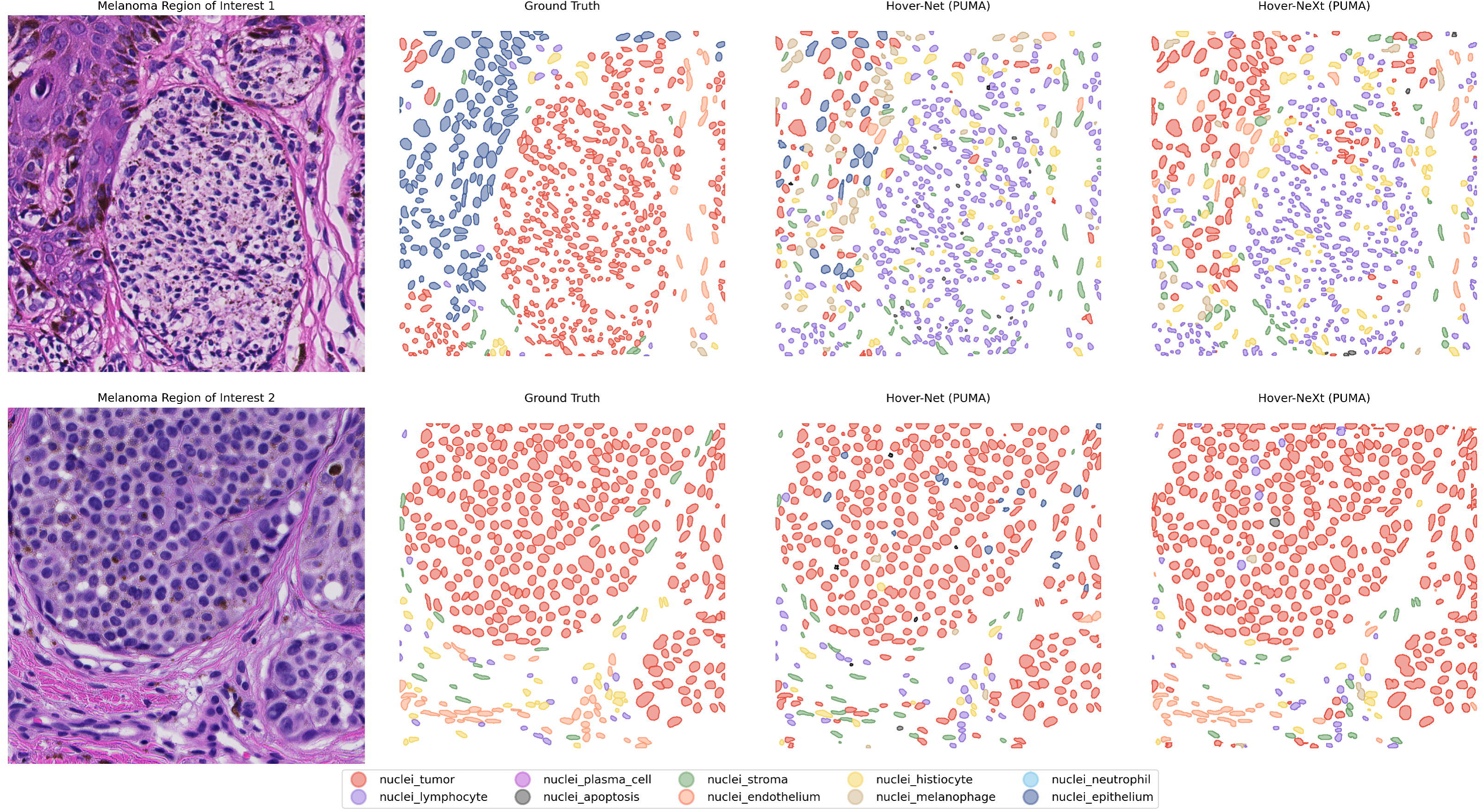
Visual results of segmentation of all nuclei classes. Results are shown for the Hover-Net and Hover-NeXt model trained on the PUMA dataset. In ROI 1 neither of the models is able to classify tumor nuclei and lymphocytes. In ROI 2 model performance is better with identification of tumor nuclei and endothelium nuclei.

#### Heuristic post processing: combining tissue and nuclei predictions

In the fourth experiment, we used the tissue mask as predicted by the nnU-Net model to improve nuclei segmentation. If a nucleus center was placed inside a tissue mask of the epidermis, the nuclei was classified as an epidermal nuclei. In addition, if a nucleus was present inside the blood vessel class, the nucleus was classified as endothelial nucleus. In Hover-Net and Hover-NeXt, the average F1 score increased from 0.27 and 0.27 to 0.31 and 0.32, respectively. This is mainly due to an increase in the F_1_ score for epithelium.

**Table 3.** F_1_ scores for heuristic post processing of the Hover-Net and Hover-NeXt output compared with their non-post processing output.

|  | Tumor | Lymphocyte | Plasma cell | Histiocyte | Melanophage | Neutrophil |
| --- | --- | --- | --- | --- | --- | --- |
| **Hover-Net (PUMA)** | 0.73 [0.72 - 0.73] | 0.67 [0.66 - 0.67] | 0.07 [0.06 - 0.09] | 0.27 [0.26 - 0.27] | 0.31 [0.29 - 0.33] | 0.15 [0.13 - 0.18] |
| **Hover-Net (PUMA - post-processed)** | 0.73 [0.73 - 0.73] | 0.67 [0.66 - 0.67] | 0.07 [0.05 - 0.08] | 0.27 [0.26 - 0.27] | 0.32 [0.30 - 0.33] | 0.15 [0.12 - 0.17] |
| **Hover-NeXt (PUMA)** | 0.70 [0.70 - 0.71] | 0.71 [0.70 - 0.71] | 0.10 [0.09 - 0.11] | 0.33 [0.32 - 0.34] | 0.26 [0.25 - 0.28] | 0.06 [0.03 - 0.08] |
| **Hover-NeXt (PUMA – post processed)** | 0.71 [0.71 - 0.71] | 0.71 [0.70 - 0.71] | 0.1 [0.09 - 0.11] | 0.33 [0.32 - 0.34] | 0.27 [0.25 - 0.28] | 0.06 [0.04 - 0.08] |
|  | Stroma | Endothelium | Epithelium | Apoptosis | Micro F<sub>1</sub> | Average F<sub>1</sub> |
| **Hover-Net (PUMA)** | 0.31 [0.29 - 0.32] | 0.15 [0.14 - 0.17] | 0.13 [0.12 - 0.14] | 0.22 [0.21 - 0.24] | 0.61 [0.61 - 0.61] | 0.27 [0.27 - 0.28] |
| **Hover-Net (PUMA - post-processed)** | 0.31 [0.29 - 0.32] | 0.2 [0.19 - 0.21] | 0.44 [0.42 - 0.46] | 0.22 [0.21 - 0.24] | 0.62 [0.62 - 0.62] | 0.31 [0.30 - 0.31] |
| **Hover-NeXt (PUMA)** | 0.21 [0.20 - 0.22] | 0.24 [0.23 - 0.25] | 0.04 [0.03 - 0.05] | 0.04 [0.04 - 0.05] | 0.61 [0.61 - 0.61] | 0.27 [0.27 - 0.27] |
| **Hover-NeXt (PUMA – post processed)** | 0.21 [0.20 - 0.22] | 0.25 [0.24 - 0.26] | 0.52 [0.50 - 0.55] | 0.05 [0.04 - 0.06] | 0.62 [0.62 - 0.62] | 0.32 [0.32 - 0.32] |

**Figure 7.**
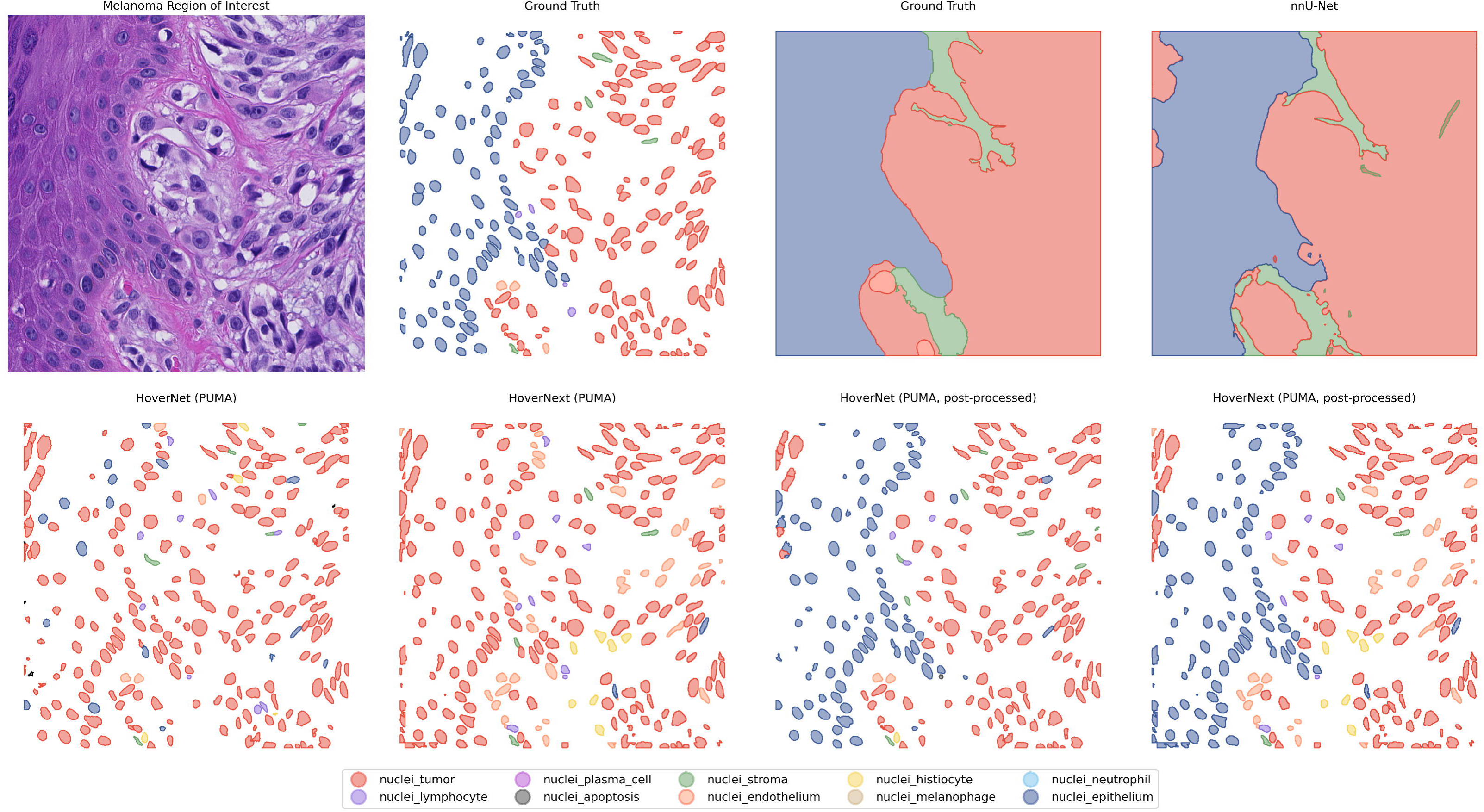
Visual results of heuristic post-processing. For clarification, the ground truth and original predictions of Hover-Net and Hover-NeXt are displayed.

## Discussion

In the present study, we show the methodology used to create the PUMA dataset, which is the first melanoma specific nuclei and tissue dataset. In addition, we provide baseline benchmarks in which we show that the Hover-NeXt model trained with three classes (tumor, lymphocyte, and other) can detect lymphocytes almost on human interobserver level performance. Finally, we show an example of the integration of tissue and nuclei annotations leading to improved nuclei segmentation. We believe that further improvement of deep learning models through this integration is possible, and therefore, the dataset will be used to organize the PUMA challenge.

Earlier studies regarding nuclei segmentation have been performed with multiple tissue datasets [17,24] or other tissue specific datasets in, for example, breast cancer [31] and colon cancer [32]. However, thus far, no melanoma specific dataset is available. Since melanocytes and melanoma cells can mimic other cell types, such as lymphocytes and stroma cells, models created with these dataset are not fully applicable for use in melanoma histopathology [13,18]. Models created with this dataset, will be usable for creation of prognostic and/or predictive biomarkers in melanoma histopathology.

The PUMA dataset combines tissue and nuclei segmentation. Earlier studies, such as the OCELOT challenge [33] and the MuTILs model [34] showed that incorporating tissue masks in nuclei prediction increased the capability of models which detect either tumor nuclei or TILs. With this dataset, the goal is to see whether implementation of tissue masks can improve multi class nuclei segmentation for classes such epidermis, endothelium, tumor, and immune cell subsets. In this paper, we demonstrated this by means of a simple post-processing technique. However, other solutions might be passing information from a tissue segmentation model directly to a nuclei segmentation model or making use of multiple sequential models [33,35]. To make use of the creativity and knowledge of the deep learning community, we initiated the PUMA challenge to assess how precise segmentation of melanoma samples can become. Participants in the PUMA challenge can choose to participate in two tracks. For track 1 the output needs to be nuclei segmentation for three classes (tumor, lymphocyte and other) and tissue segmentation for all classes whereas in track 2 participants need to segment all nuclei classes and tissue segmentation for all classes.

With the dataset, we also aimed to assess the performance of existing models usable in skin and/or melanoma histopathology. Due to unavailability, we were unable to assess the LUNIT scope IO model. However, we were able to evaluate the NN192 algorithm, which is used in multiple studies in which lymphocytes in melanoma histopathology were associated with a decreased chance of recurrence [36] and an increased chance of responding to immune checkpoint inhibition therapy [7]. Surprisingly, the NN192 model showed the lowest capability to detect lymphocytes. This might be due to variations in staining and scanners used in the studies performed earlier and the inability of the techniques used to correct for this. The Hover-Net and Hover-NeXt algorithm architectures can compensate for this by extensive data augmentation in the training (Hover-Net, Hover-NeXt) and inference stages (Hover-NeXt), accounting for better results with the out of the box model trained on the PanNuke dataset. However, both models trained on the PanNuke dataset still showed diminished performance in more difficult to segment melanoma samples when compared to the Hover-NeXt model trained on the PUMA dataset itself.

A strength of this study is the annotation process by a medical expert and an expert dermatopathologist using context ROI to more accurately segment and classify nuclei. In addition, we validated the annotations with a second independent pathologist (GB). From this, it became clear that there is a low interobserver agreement in the less common classes, such as histiocytes, melanophages, and stroma. Melanophages are a subtype of histocytes that have phagocytosed melanin, resulting in a more pigmented appearance. However, there is a continuum between the cells, which could explain the difference in classification. Stroma cells, in addition, are present just like histiocytes in the tissue around the tumor, and it is not always possible to distinguish them from histiocytes on H&E stained slides.

A second strong point of the study is the manual selection of ROIs. This allowed us to focus on immune cell subsets and less common nuclei and tissue classes. In addition, we tried to include regions with artifacts or much pigmentation. We believe this makes the dataset and the models more applicable to whole slide images, which by nature have artifacts, unsharp regions, and, especially in the case of melanoma, pigmentated more difficult to segment regions.

A downside of the dataset and the study is that all samples are, due to privacy regulations, from one scanner and a single hospital. However, 76 out of 310 cases are consultation cases from referral hospitals already leading to substantial staining variation. In addition, we believe that through stain normalization and data augmentation, models can become less sensible to domain-adversarial effects. This makes models trained on this dataset applicable to whole slide images from other hospitals and scanner types.

Our study did not demonstrate high performance in segmentation of all nuclei classes. This is partly due to the used models losing discriminative ability by introducing more classes. In addition, some classes are less common in the dataset such as apoptosis or histiocytes. Furthermore, there is overlap between classes such as melanophages and histiocytes. We believe that further improvement of models is possible and hope to that through leveraging the knowledge of the deep learning community in the PUMA challenge this will become more clear.

In conclusion, we created a novel dataset with tissue and nuclei segmentations in advanced cutaneous melanoma. With this dataset we showed that deep learning based lymphocyte segmentation can achieve performance levels close to those of human interobserver agreement. In addition, we demonstrated that it is possible to more accurately segment nuclei classes by incorporating tissue predictions. However, we believe that further improvement is possible by further integration of tissue and nuclei segmentation. Future work in the PUMA challenge will demonstrate to what extent segmentation of different nuclei can be improved.

### Potential implications

The PUMA dataset and the deep learning models created with this dataset can be used for biomarker generation in the treatment of melanoma. Thus far, it is known that lymphocytes have predictive value for melanoma recurrence and immune checkpoint inhibition treatment response [6,7,37]. However, this is done either by manual scoring or scoring by automated models, which are not generalizable to new unseen tissue and scanners. With this public dataset and the generalizable deep learning models developed from it, the next step will be to translate these models into clinical application. This will result in more personalized treatment plans, such as de-escalation of immune checkpoint inhibition therapy in patients with advanced melanoma or a less intensive follow-up regimen in patients with primary melanoma. Next to the assessment of TILs as an explainable biomarker, this study will also enable future biomarker discovery studies through the assessment of other nuclei and tissue types.

In addition, this study adds a new step towards further improvement of nuclei segmentation deep learning models in H&E stained histological slides by combining tissue segmentation models into multiclass nuclei segmentation models. This process is closer to how a pathologist evaluates histological slides and already resulted in improved nuclei segmentation benchmark scores.

## Methods

### Dataset generation

For the dataset, digitized melanoma whole slide H&E stained images generated through regular diagnostic procedures were used from the archive of the UMC Utrecht. All slides were scanned with a Nanozoomer XR C12000–21/− 22 (Hamamatsu Photonics, Hamamatsu, Shizuoka, Japan) at 40× magnification with a resolution of 0.23 µm per pixel. Out of 310 cases in the dataset, 76 are consultation cases from referral hospitals or general practitioners leading to variation in used staining protocols. From each slide, a 40× magnified ROI of 1024×1024 pixels was selected for annotation. In addition, a context ROI of 5120×5120 pixels was sampled to provide information about the broader context for the annotation process. Selection was done by a trained medical expert (M.S.) and subsequently verified by an expert dermatopathologist (W.B.). Manual ROI selection ensured diverse tissue and nuclei types and the inclusion of more difficult to segment areas due to blurring, pigmentation, and scanning artifacts.

Nuclei segmentations were generated with Hover-Net pretrained on the PanNuke dataset [16,17]. Manual annotation adjustment was performed by author M.S. using Qupath with the following nuclei categories: tumor, stroma, vascular endothelium, histiocyte, melanophage, lymphocyte, plasma cell, neutrophil, apoptotic and epithelium [26]. Annotation categories were based on earlier datasets. In addition, we chose categories based on possible predictive value. All annotations were checked and corrected where needed by a dermatopathologist (W.B.). Intra- and inter-observer agreement (by pathologist G.B.) were determined on 12 randomly selected ROIs.

Tissue segmentations were created manually with QuPath by author M.S. using the following categories: tumor, stroma, epidermis, necrosis, blood vessel, and background. Annotation categories are based on the approach used by Hwang et al., who segmented TILs in tumor and stroma areas, and the guidelines from the International Immuno-Oncology Biomarker Working Group [15,38]. Annotations are checked and corrected when needed by a dermatopathologist (W.B.). Intra- and inter-observer agreement (by pathologist G.B.) were determined on 12 randomly selected ROIs to ensure the inclusion of all tissue types.

### Benchmark models

#### Nuclei segmentation

To establish baseline nuclei segmentation benchmarks, we performed nuclei segmentation for two sets of experiments. The first experiment compares models that output three nuclei categories: tumor, lymphocyte, and other. The second experiment is an analysis of the segmentation of all individual nuclei categories. Model training was performed with 5-fold cross validation on the public training dataset without adjusting training parameters or data augmentation of the algorithm used. Inference was performed on the 94 ROIs of the final hidden test set of the PUMA challenge. The remaining 10 ROIs will be used in the PUMA challenge for sanity checking of submitted models.

For the first experiment, the following models were compared: NN192, Hover-Net trained on the PanNuke dataset, Hover-Net trained on the PUMA dataset, Hover-Next trained on the PanNuke dataset, Hover-Next trained on the PUMA dataset [16,17,37,39]. The NN192 model outputs four categories: tumor, lymphocytes, stroma, and other. From this output, the stroma and other categories were merged into one other category. The models trained on the PanNuke dataset classify nuclei into 5 categories: neoplastic, non-neoplastic epithelial, inflammatory, connective, and apoptotic. For the benchmark comparison, neoplastic and inflammatory nuclei were used; non-neoplastic epithelial, connective, and apoptotic were merged into the other category. For training on the PUMA dataset, the plasma cell and lymphocyte categories were merged into a single lymphocyte category, and the remaining classes were combined into the other category. For the second experiment, Hover-Net and Hover-NeXt were trained on all samples and compared.

For the calculation of the F1 score, the center distance between the predicted nuclei and the ground truth nuclei was used. For each ground truth nucleus, predictions within a 15 pixel (3.3 μm) were identified. This radius is smaller than the average size of lymphocytes, which form the smallest nuclei in the dataset. Matching was performed based on the highest predictive score (if available) or the shortest distance. After matching, the ground truth was censored until all ground truth nuclei were either matched or classified as a false negative. Using the identified true positives, false positives, and false negatives, precision (all correct predictions divided by all predictions) and recall (all correct predictions divided by all ground truth nuclei) were calculated. The class F1 score was computed as the harmonic mean of precision and recall, ranging from 0 to 1. Finally to compare models, micro F_1_ (aggregation of TP, FP, and FN over all classes, followed by F1 score calculation) and Average F_1_ (the average of class F_1_ scores) were calculated [40]. Results are shown with a 95% confidence interval which is calculated through bootstrapping the samples.

#### Tissue Segmentation

For tissue segmentation, nnU-Net and Mask2Former were used to establish baseline benchmarks [28,41]. For training of nnU-Net the same fivefold cross validation was used to create an ensemble model. Mask2Former was pretrained on the COCO instance segmentation task using a Swin Transformer backbone. Images were resized to 512 × 512 pixels before loading into the model. For our experiment we replaced the backbone with the UNI pathology foundation model, which is better able at feature extraction from H&E stained histopathology, after which we finetuned the model on the whole training dataset [29,30,42]. Both models were used for inference on the final hidden test set from the PUMA challenge.

Both models and intra- and interobserver agreement were evaluated using the DICE score. The DICE score is a harmonic mean between 0 and 1, in which 1 is a perfect segmentation prediction, and 0 is no correct prediction. This can result in inflated high average DICE scores if a tissue class is only present in a few samples, as the DICE score is 1 in the case of a correct absent prediction. To accommodate this, we calculated not only the average DICE score over all samples but also the micro average DICE. This is the DICE score for all predictions concatenated along one axis, resulting in a prediction mask of 1024 × 96.256 pixels. Results are shown with a 95% confidence interval which is generated through bootstrapping the sample results.

#### Post-processing

For post-processing nuclei, centers were calculated using the Point function from the python Shapely library. From these points, the spatial location inside a tissue mask prediction was determined by using the GeoPandas Python Package. Based on this location the classification label of the nuclei was adjusted.

## Data Availability

Part of the data produced are available online, the rest is available on reasonable request.

https://zenodo.org/records/13859989

https://puma.grand-challenge.org/puma/

https://github.com/tueimage/PUMA-challenge-eval-track1

https://github.com/tueimage/PUMA-challenge-eval-track2

https://github.com/tueimage/PUMA-challenge-baseline-track1

https://github.com/tueimage/PUMA-challenge-baseline-track2

## Availability of source code and requirements

Project name: PUMA

Project home page: https://pumachallenge.com/

Operating system: Platform independent

Programming language: Python 3.10

Other requirements: Hover-Net (available at: https://github.com/vqdang/hover_net), Hover-NeXt (https://github.com/digitalpathologybern/hover_next_train and https://github.com/digitalpathologybern/hover_next_inference).

nnU-Net (available at: https://github.com/MIC-DKFZ/nnUNet) and Mask2Former (available at https://huggingface.co/docs/transformers/en/model_doc/mask2former).

QuPath 0.5.0 (available at https://qupath.github.io/)

The NN192 classification algorithm (available at https://github.com/acsbal/Automated-TIL-scoring-

QuPath-Classifier-for-Melanoma) and QuPath 0.1.2 (https://github.com/qupath/qupath/releases/tag/v0.1.2).

The code and weights for the PUMA challenge inference baseline solutions and metric calculation can be found here: (https://zenodo.org/records/13897135, https://github.com/tueimage/PUMA-challenge-eval-track1, https://github.com/tueimage/PUMA-challenge-eval-track2, https://github.com/tueimage/PUMA-challenge-baseline-track1, https://github.com/tueimage/PUMA-challenge-baseline-track2)

No changes were made to the training code for the used algorithms.

License: The PUMA codebase is licensed with a CC0 1.0 license (dataset) and the MIT license.

Restrictions to use by non-academics: Both the CC0 1.0 license (dataset) and the MIT license (codebase) allow for non-commercial use. License terms can be reviewed for details.

## Data Availability

The PUMA training dataset is available on Zenodo at: https://zenodo.org/records/10940194.

The PUMA test dataset is available on reasonable request.

## List of abbreviations

TILs: Tumor infiltrating lymphocytes
H&E: Hematoxylin and Eosin
ROI: Region of interest

## Declaration

The Biobank Research Ethics Committee (TCBio) UMC Utrecht confirms that it has reviewed the release file in accordance with the UMC Utrecht Biobank Regulations and all other applicable regulations and laws. Based on the requirements as defined in these regulations and laws, the TCBio UMC Utrecht hereby issues an approval of the aforementioned dataset (reference number TCBio 23-270/U-B).

## Competing interests

The authors declare the following financial interests/personal relationships which may be considered as potential competing interests:

Karijn P.M. Suijkerbuijk reports a consulting/advisory relationship with Abbvie and Sairopa. She received honoraria from Bristol Myers Squibb and research funding from TigaTx, Bristol Myers Squibb, Philips, Genmab and Pierre Fabre. All paid to institution

The remaining authors of this manuscript have no conflicts of interest to disclose.

## Funding

This research was funded by an unrestricted grant of Stichting Hanarth Fonds, The Netherlands..

## Authors’ contributions

Conceptualization: Schuiveling, Blokx, Suijkerbuijk, Veta

Methodology: Schuiveling, Blokx, Veta, Liu, Breimer

Software: Schuiveling, Eek, Liu

Investigation: Schuiveling, Liu

Resources: Blokx, Suijkerbuijk, Veta

Data Curation: Schuiveling, Blokx, Breimer

Writing, Original Draft: Schuiveling, Liu

Writing, Review & Editing: Blokx, Suijkerbuijk, Veta, Eek, Breimer

Supervision: Blokx, Suijkerbuijk, Veta

Project administration: Blokx, Suijkerbuijk, Veta

Funding acquisition: Suijkerbuijk, Blokx, Veta

